# Arterial Stiffness, Smoking Status, and Cardiovascular Mortality in Asymptomatic U.S. Adults

**DOI:** 10.64898/2026.02.09.26345932

**Authors:** Patrick Cheon, Mohamed Mostafa, Alexander Grdzelishvili, David Cornea, Jessica Liu, Richard Kazibwe

## Abstract

**Objective:** To examine whether the association between smoking status and cardiovascular (CV) mortality differs by arterial stiffness, assessed by pulse pressure index (PPI), among U.S. adults without baseline cardiovascular disease (CVD).

**Methods:** Using data from the National Health and Nutrition Examination Survey (NHANES) 2005–2016, we analyzed 16,605 adults aged 40–79 years without baseline CVD, with mortality follow-up through December 31, 2019. PPI was calculated as (systolic blood pressure [SBP] − diastolic blood pressure [DBP])/SBP and split at the cohort median (0.415) as low versus high. Smoking status was classified as never, former, or current, yielding six joint PPI–smoking groups. Cox models estimated hazard ratios (HRs) and 95% confidence intervals (CIs) for CV mortality, adjusting for demographics and cardiometabolic risk factors.

**Results:** Over a median follow-up of 8.4 years, 518 CV deaths (3.1%) occurred. Among individuals with low PPI, former smokers had CV mortality comparable to never smokers (HR 0.86, 95% CI 0.56–1.33), whereas current smokers remained at elevated risk (HR 2.51, 95% CI 1.65–3.81). This pattern was not observed in the high PPI stratum, where both former and current smokers had significantly higher CV mortality than never smokers.

**Conclusion:** Former smokers with low PPI had CV mortality similar to never smokers, whereas former smokers with high PPI remained at elevated risk. These findings suggest that the CV benefit of smoking cessation may be greatest when arterial stiffness is minimal, supporting early cessation before substantial vascular aging occurs.

## Background

Cardiovascular disease (CVD) remains a leading cause of mortality worldwide, highlighting the need for improved strategies to identify high-risk individuals for targeted interventions [1]. Cigarette smoking is a major modifiable determinant of cardiovascular (CV) risk and continues to contribute substantially to preventable morbidity and mortality [2-5]. Although smoking cessation is known to reduce CV risk, the degree of risk reduction varies considerably across individuals [4, 6, 7]. In particular, the factors that modify the CV benefits of cessation are not fully understood. One potential factor is the extent of vascular aging, which may influence how fully CV risk normalizes after quitting [8, 9].

Arterial stiffness, a marker of vascular aging, has emerged as a strong predictor of CV events and mortality in population-based studies, independent of traditional risk factors [10-13]. Arterial stiffness reflects long-term exposure to adverse cardiometabolic and lifestyle factors and may provide important prognostic information in preventive settings [14]. Accordingly, practical measures that capture arterial stiffness may help operationalize vascular aging for risk stratification in both clinical and population-based research.

The pulse pressure index (PPI), calculated as the ratio of pulse pressure to systolic blood pressure (SBP), is a simple measure of arterial stiffness [15]. Compared with pulse pressure alone, PPI may better capture relative pulsatile burden while reducing confounding by overall blood pressure (BP) level [16-18]. Importantly, PPI is easily obtainable in large-scale surveys and clinical practice, making it a potentially useful marker for population-level CV risk stratification [15, 17].

Despite established independent associations of smoking and arterial stiffness with adverse CV outcomes, these factors have only been examined separately. Consequently, it remains unclear whether the association between smoking status and CV mortality differs according to the degree of arterial stiffness. Such findings could reveal whether markers of arterial stiffness can help identify subgroups in whom smoking cessation may confer greater long-term benefit.

Therefore, we used data from the National Health and Nutrition Examination Survey (NHANES) 2005-2016 to examine the individual and joint associations of PPI and smoking status with CV mortality among U.S. adults without baseline CVD.

## Methods

### Study Population

NHANES is a program designed to assess health and nutritional status among U.S. adults and children living in the community. It is conducted by the National Center for Health Statistics (NCHS), part of the Centers for Disease Control and Prevention. This analysis used data from NHANES cycles 2005-2016. All participants gave written informed consent, and the study protocol was approved by the NCHS institutional review board. Further information on the survey’s design, methodology, and data availability has been documented in prior publications [19-21].

For the purpose of this analysis, we included NHANES participants aged 40-79 years with available BP and mortality data at baseline. We excluded those with prior CVD (coronary heart disease, myocardial infarction, angina, stroke, or congestive heart failure), those with missing or implausible BP values (SBP <70 or >250 mmHg; diastolic blood pressure (DBP) <40 or >150 mmHg), those with missing smoking status, missing mortality follow-up, and missing key covariates needed for analysis. After all exclusions, 16,605 participants were included in the analysis (Figure 1).

**Fig. 1.**
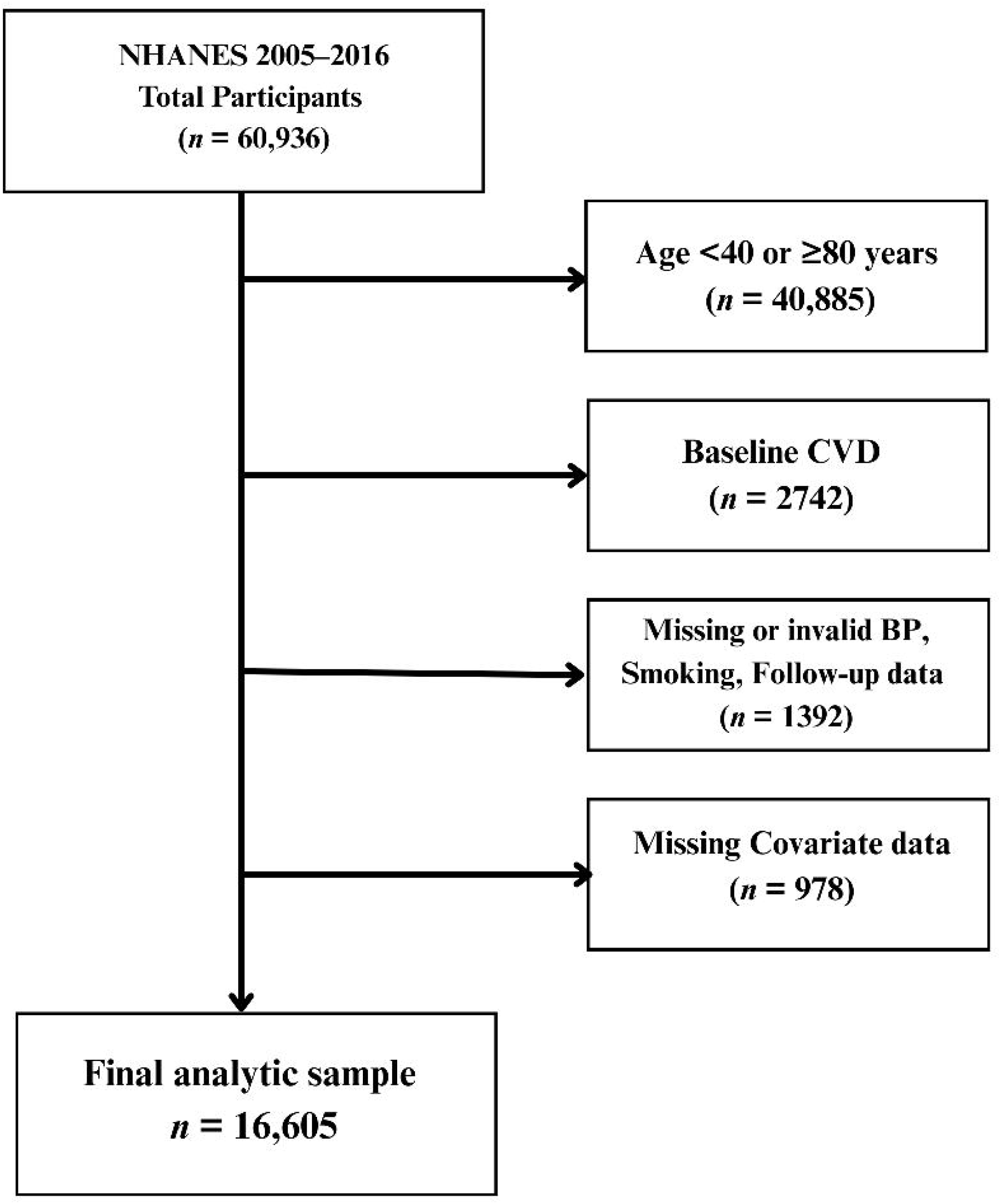

### Ascertainment of Arterial Stiffness

During NHANES assessments at mobile examination centers, BP was measured by trained staff using a mercury sphygmomanometer after a 5-minute seated rest, and up to three readings were averaged. Arterial stiffness was assessed using the pulse pressure index (PPI), calculated as [(SBP − DBP) ÷ SBP]. Participants were categorized into two groups according to the median PPI value (0.415): low PPI (≤0.415) and high PPI (>0.415), consistent with previous studies examining pulse pressure [22, 23].

### Defining Smoking Status and Analysis Groups

Smoking status was ascertained via self-report during household interviews. Participants were classified as never, former, or current smokers based on standardized questionnaire responses. To evaluate the combined effects of smoking and arterial stiffness, participants were additionally classified into six groups based on the cross-classification of PPI category and smoking status: low PPI with never smoker, low PPI with former smoker, low PPI with current smoker, high PPI with never smoker, high PPI with former smoker, and high PPI with current smoker.

### Ascertainment of Cardiovascular Mortality

The primary outcome was CV mortality, determined through linkage to the National Death Index with follow-up through December 31, 2019. Vital status and cause of death were obtained from NHANES public-use linked mortality files created by NCHS using a validated probabilistic matching algorithm. CV mortality was identified using International Classification of Diseases, 10th Revision (ICD-10) codes I00-I78 corresponding to underlying causes of death. Person-years were calculated from the baseline examination date until date of death or end of follow-up, whichever occurred first.

### Other Variables

Demographics (age, sex, race/ethnicity) and education level were self-reported during household interviews. Race/ethnicity was categorized as non-Hispanic White, non-Hispanic Black, Mexican American, Other Hispanic, or Other race. Education was assessed as years of schooling completed. Body mass index (BMI) was calculated as weight in kilograms divided by height in meters squared using data from a physical examination conducted at a mobile examination center. Diabetes and kidney failure were determined from self-reported questionnaire data. Total cholesterol was measured using standardized enzymatic assays, and statin use was ascertained from prescription medication questionnaires, as reported by the NCHS [24].

### Statistical Analysis

We compared participant characteristics across the six PPI–smoking groups using chi-square tests for categorical variables. Continuous variables were summarized as means with standard deviations (SD) and compared using one-way analysis of variance. Categorical variables are presented as counts and percentages.

We calculated CV mortality incidence rates per 1000 person-years stratified by PPI levels and smoking status. Multivariable Cox proportional hazards models were used to assess the associations of PPI and smoking status, separately and in combination, with CV mortality. PPI was modeled both continuously (per SD increase) and categorically (low vs. high), and smoking status was included using indicator variables. We first evaluated main effects of PPI and smoking, then modeled the six joint PPI–smoking groups, using low PPI/never smoker as the reference category. Models were adjusted as follows: Model 1 was adjusted for sociodemographic variables: age, sex, race/ethnicity, education, and income. Model 2 was further adjusted for traditional CV risk factors, including BMI, diabetes, kidney failure, total cholesterol, and statin use. Multiplicative interaction between PPI and smoking status was assessed by including cross-product terms and comparing nested models with likelihood ratio tests.

Kaplan-Meier curves for CV mortality were generated by smoking status within low and high PPI strata, with corresponding numbers at risk. We estimated 5- and 10-year cumulative incidence of CV mortality for each of the six PPI–smoking groups and displayed these estimates with 95% confidence intervals (CI).

All statistical analyses were performed using R software (version 4.3.0; https://www.r-project.org) (accessed on 5 February 2025). A two-sided α of 0.05 was used for hypothesis testing.

## Results

### Baseline Characteristics

The mean age of the cohort was 56.5 years, 51.9% were women, and 41.2% were non-Hispanic White. Over a median follow-up of 8.4 years (interquartile range 5.7–11.2), 518 CV deaths occurred (3.1%). Participants with high PPI were older, had higher SBP and wider pulse pressure, and more often had hypertension, diabetes, kidney disease, and statin use than those with low PPI. Current smokers tended to have lower BMI and lower educational attainment and were more likely to report low income. The most adverse cardiometabolic profile was observed among participants with high PPI who smoked (either former or current); they had the greatest burden of hypertension and diabetes and the highest rates of CV mortality (Table 1).

**Table 1.**
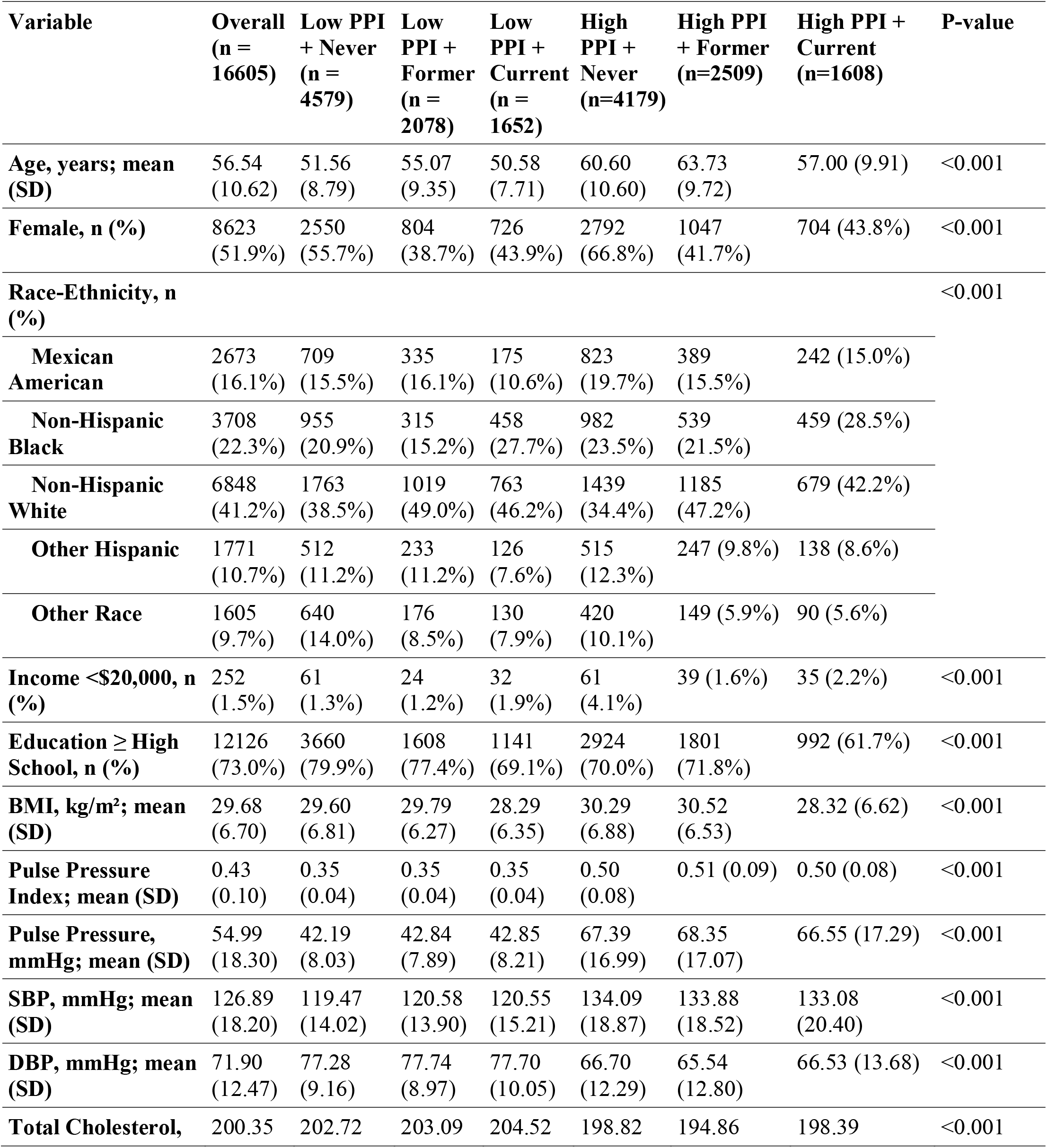
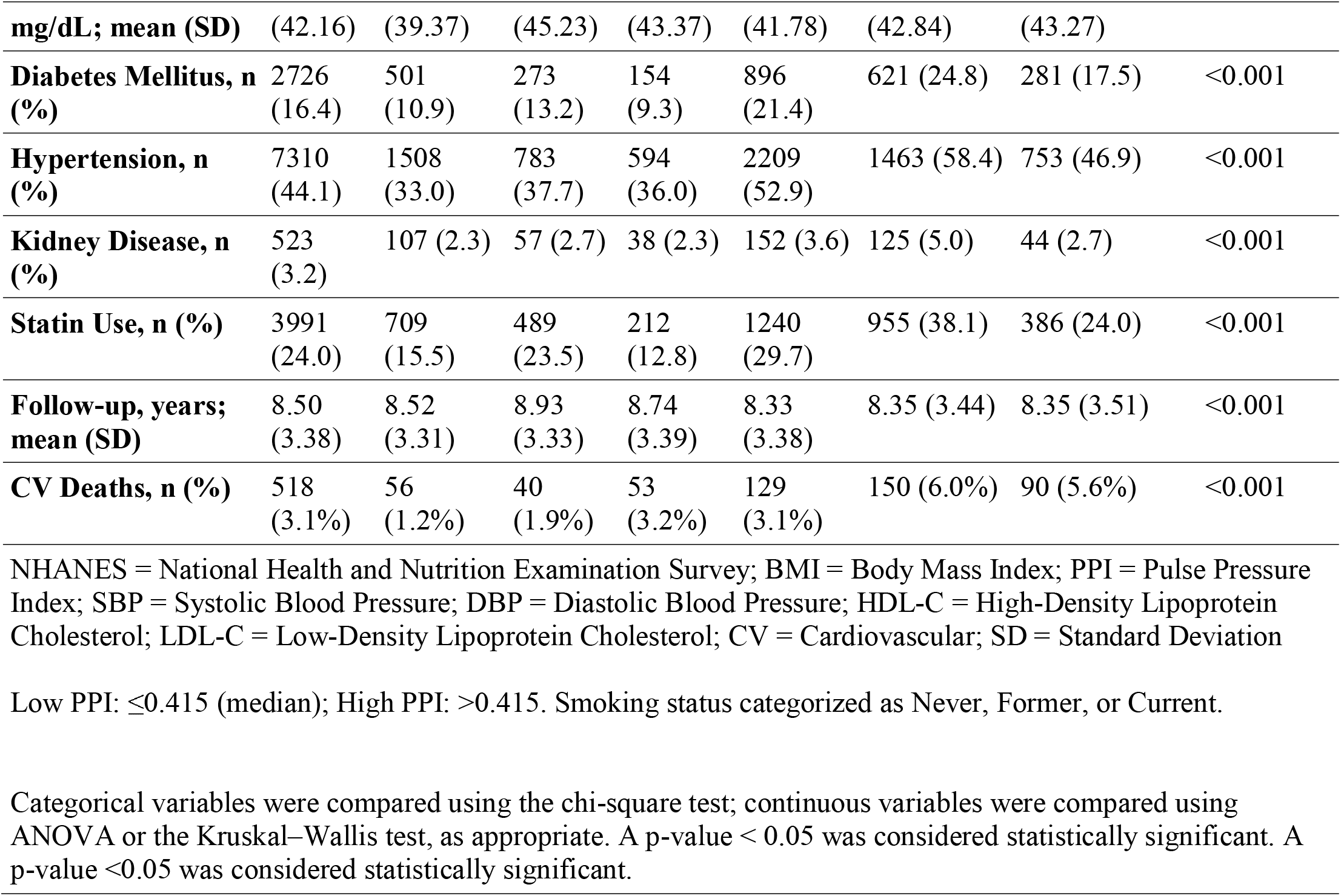
Baseline Characteristics of Study Population according to PPI and smoking status group in NHANES 2005-2016.

### Joint Association of PPI and Smoking Status with CV Mortality

Among individuals with low PPI, former smokers had CV mortality risk similar to never smokers. In contrast, current smokers with low PPI had more than double the risk. In the high PPI group, both former and current smokers had higher CV mortality than never smokers, with the greatest risk seen in current smokers (Table 2). The interaction hazard ratio (HR) comparing the smoking effect in high versus low PPI strata was 1.66 (95% CI 1.01–2.71; P = 0.044) for former smokers, whereas no clear interaction was seen for current smokers (HR 0.97, 95% CI 0.60–1.59; P = 0.907). When PPI was modeled as a continuous variable, interaction terms with smoking were not statistically significant (PPI × former: HR 3.38, 95% CI 0.54–21.13; P = 0.193; PPI × current: HR 0.47, 95% CI 0.05–4.10; P = 0.496).

**Table 2.** Association of Combined Pulse Pressure Index and Smoking Status with Cardiovascular Mortality: NHANES 2005-2016.

| PPI Status | Smoking Status | No. Events (%) | Rate Per 1,000 | Model 1 |  | Model 2 |  |
| --- | --- | --- | --- | --- | --- | --- | --- |
|  |  |  |  | HR (95% CI) | P-Value | HR (95% CI) | P-Value |
| Low PPI | Never | 55 (1.2%) | 1.40 | Ref. | --- | Ref. | --- |
|  | Former | 40 (1.9%) | 2.20 | 0.87 (0.57-1.32) | 0.520 | 0.86 (0.56-1.33) | 0.507 |
|  | Current | 53 (3.2%) | 3.70 | 2.45 (1.66-3.61) | <0.001 | 2.51 (1.65-3.81) | <0.001 |
| High PPI | Never | 130 (3.1%) | 3.70 | Ref. | --- | Ref. | --- |
|  | Former | 150 (6.0%) | 7.20 | 1.41 (1.11-1.80) | 0.006 | 1.47 (1.14-1.89) | 0.003 |
|  | Current | 90 (5.6%) | 6.70 | 2.40 (1.81-3.18) | <0.001 | 2.75 (2.05-3.70) | <0.001 |

*PPI = pulse pressure index; HR = hazard ratio; CI = confidence interval*
*Model 1: Adjusted for age, sex, race/ethnicity, education*
*Model 2: Adjusted for age, sex, race/ethnicity, education, body mass index, diabetes, kidney disease, total cholesterol, and statin use*

### Survival Curves

Kaplan-Meier curves stratified by PPI level (Figure 2) further illustrated survival patterns. Among participants with low PPI, never smokers had the most favorable survival, former smokers showed a modestly steeper decline, and current smokers experienced the steepest decline over follow-up. In the high PPI group, overall survival was lower across all smoking categories. Never smokers had the highest survival. Curves for former and current smokers were closer together and declined more rapidly than those of never smokers.

**Fig. 2.**
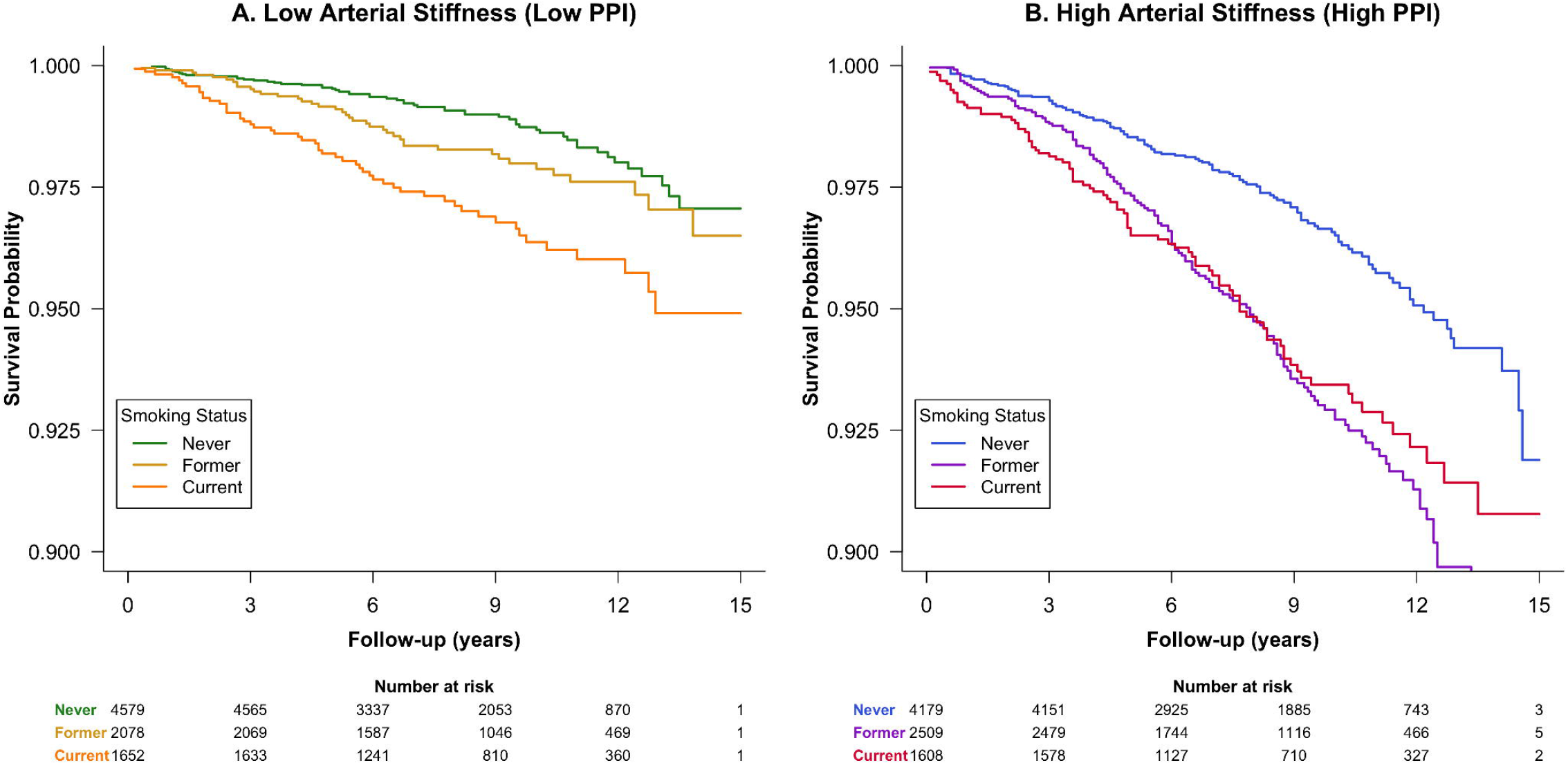

### Absolute Risk

Ten-year absolute risks of CV mortality estimated from Kaplan-Meier curves ranged from 1.3% (95% CI 0.9–1.7%) in low PPI/never smokers to 7.3% (95% CI 6.0–8.5%) in high PPI/former smokers. In the low PPI group, former smokers had only a modest increase in 10-year risk compared with never smokers (2.1% vs. 1.3%), whereas in the high PPI group, former smokers had substantially higher risk than never smokers (7.3% vs. 3.5%). Corresponding numbers needed to treat with smoking cessation over 10 years were 125 for individuals with low PPI and 26 for those with high PPI (Figure 3).

**Fig. 3.**
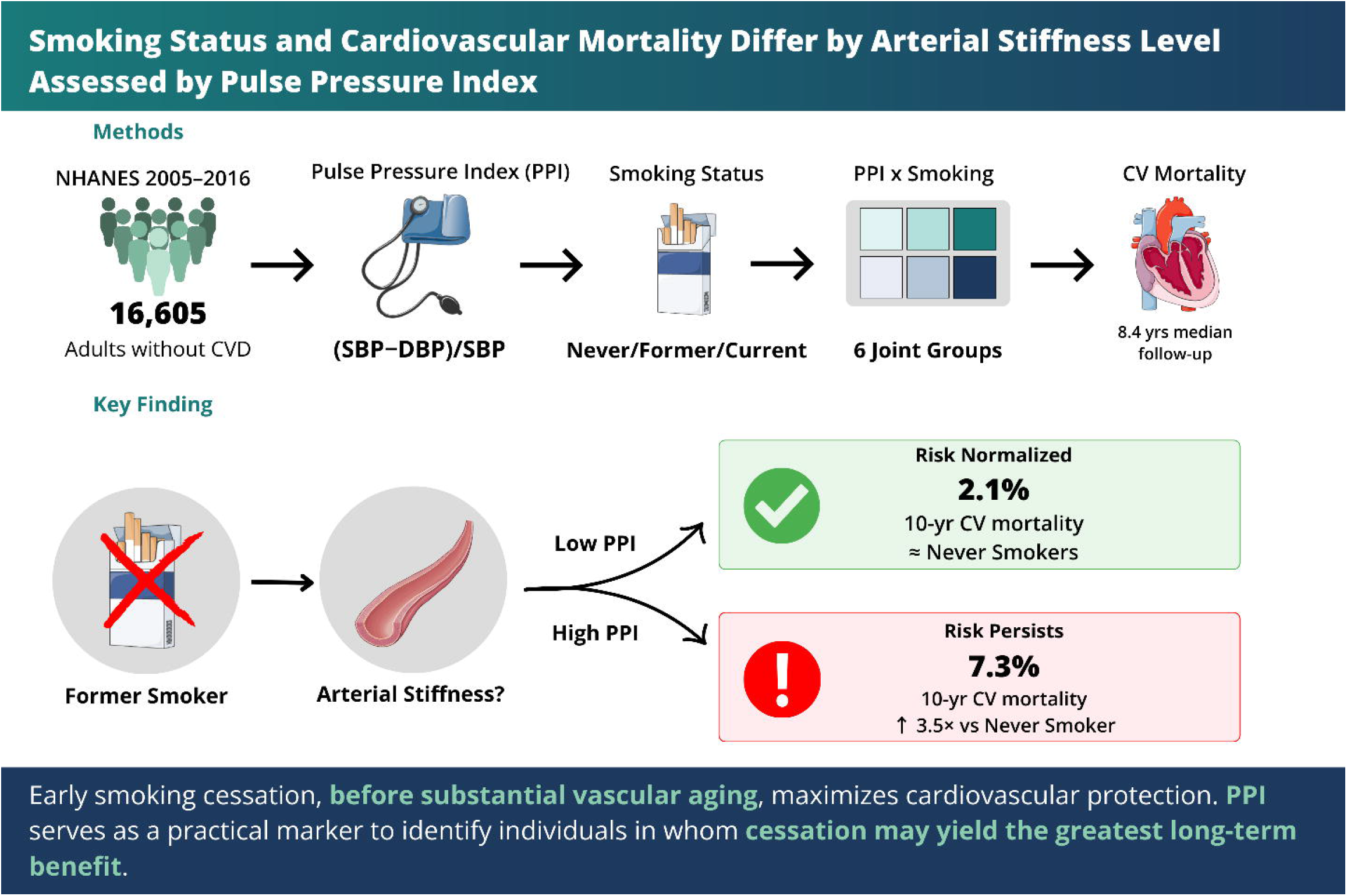

## Discussion

In this nationally representative cohort of U.S. adults without baseline CVD, the CV consequences of smoking differed according to the degree of arterial stiffness. Among individuals with low PPI, former smokers had CV mortality similar to never smokers, suggesting that cessation may largely reverse CV risk when vascular aging is minimal. Among those with high PPI, both former and current smokers had elevated CV mortality, and the gap between them was narrower. This pattern suggests that once substantial arterial stiffness has developed, the CV benefits of smoking cessation may be attenuated.

Notably, among individuals with high PPI, former smokers had slightly higher absolute 10-year CV mortality than current smokers (7.3% vs. 6.6%), though confidence intervals overlapped substantially. This counterintuitive finding may reflect the well-described “sick quitter” phenomenon, wherein individuals often quit smoking following a health event or new diagnosis [25, 26]. This would result in a former smoker population with a greater proportion of higher-risk individuals at baseline. Additionally, current smokers who survive into older age without CVD may represent a hardier subset with greater resilience to smoking-related harm [27]. Importantly, this difference was not statistically significant, and the overall pattern supports the primary conclusion that smoking cessation yields attenuated CV benefit once substantial arterial stiffness has developed.

Prior studies have shown that smoking accelerates arterial stiffening through oxidative stress, endothelial dysfunction, and chronic inflammation [28-30]. Longitudinal evidence suggests that cessation may partially reverse these changes [8, 9]. Our findings extend this work by demonstrating that CV mortality risk associated with smoking varies by the degree of arterial stiffness. Notably, former smokers experienced near-normalization of risk only when vascular aging was minimal.

### Clinical Implications

Our findings have several implications for CV risk assessment and prevention. First, the observation that former smokers with low PPI had CV mortality comparable to never smokers supports the benefit of early smoking cessation, particularly before the development of substantial vascular aging. This reinforces public health messaging that quitting smoking at any age is beneficial, but that earlier cessation may yield greater long-term CV protection.

Second, PPI may serve as a practical, readily obtainable marker to identify individuals in whom smoking cessation is likely to confer the greatest benefit. The substantial difference in numbers needed to treat (125 for individuals with low PPI versus 26 for those with high PPI) highlights the potential utility of incorporating arterial stiffness assessment into smoking cessation counseling. Individuals with evidence of advanced vascular aging may warrant more intensive risk factor modification beyond smoking cessation alone.

Third, the attenuation of differences between former and current smokers in the high PPI group emphasizes the importance of primary prevention. Once substantial arterial stiffness is present, overall CV risk is elevated regardless of subsequent behavioral modification, highlighting that prevention efforts should target individuals before irreversible vascular changes have occurred.

### Strengths and Limitations

Our study has several strengths. First, we used data from NHANES, a rigorously conducted national survey with standardized protocols for BP measurement, covariate ascertainment, and mortality linkage, enhancing internal validity and generalizability to the U.S. adult population. Second, by focusing on adults without baseline CVD, we were able to examine primary prevention-relevant associations free from reverse causation by prevalent disease. Third, the joint classification by PPI and smoking status allowed us to move beyond single-exposure analyses and evaluate clinically meaningful risk phenotypes. Fourth, the use of both relative and absolute risk metrics, including 10-year cumulative incidence and numbers needed to treat, provides actionable information for clinical decision-making. Finally, the follow-up period (8.4 years) provided sufficient events to detect meaningful associations with CV mortality.

This study also has limitations. First, as an observational analysis, residual confounding cannot be excluded despite multivariable adjustment for established CV risk factors. Second, smoking status was self-reported and may be subject to recall or social desirability bias; additionally, we lacked detailed data on cumulative pack-years or duration since cessation, which could further clarify risk gradients among former smokers and inform the timeline over which cessation benefits accrue. Third, although NHANES employs complex sampling to achieve national representativeness, the majority of participants were non-Hispanic White, and participants classified as “other” race/ethnicity were heterogeneous, which may limit generalizability to specific racial and ethnic subgroups. Finally, we used PPI as a surrogate for arterial stiffness; direct measures such as carotid-femoral pulse wave velocity were not available, and future studies should examine whether these measures provide complementary prognostic information.

## Conclusions

In a large, nationally representative cohort of U.S. adults free of baseline CVD, individuals who were current smokers and had high PPI experienced the greatest burden of CV mortality. Our findings also suggest that the CV benefit of smoking cessation may depend on the degree of arterial stiffness. Former smokers with low arterial stiffness had mortality risk similar to never smokers, whereas former smokers with high arterial stiffness remained at elevated risk. These results support PPI as a practical marker of vascular aging that can refine risk stratification among people with a history of smoking.

## Data Availability

The data analyzed in this study are publicly available from the National Health and Nutrition Examination Survey (NHANES), conducted by the Centers for Disease Control and Prevention (CDC). NHANES datasets, including laboratory, examination, and questionnaire, can be accessed at https://www.cdc.gov/nchs/nhanes/. Linked mortality follow-up data are available through the National Center for Health Statistics (NCHS) Linked Mortality Files. No new data were created for this study. Derived variables and analytic code used to generate the results are available from the corresponding author upon reasonable request.

https://www.cdc.gov/nchs/data-linkage/mortality-public.htm

## List of Abbreviations

BMI: Body mass index
BP: Blood pressure
CI: Confidence interval
CV: Cardiovascular
CVD: Cardiovascular disease
DBP: Diastolic blood pressure
HR: Hazard ratio
ICD-10: International Classification of Diseases, 10th Revision
NCHS: National Center for Health Statistics
NHANES: National Health and Nutrition Examination Survey
PPI: Pulse pressure index
SBP: Systolic blood pressure
SD: Standard deviation

## Declarations

### Authors’ contributions

Conceptualization: PC, RK. Data curation: PC. Formal analysis: PC. Methodology: PC, RK. Project administration: PC. Visualization: PC. Writing - original draft: PC, DC, AG. Writing - review & editing: MM, JL, RK.

### Ethics approval and consent to participate

Ethical review and approval were waived for this study because it involved secondary analysis of publicly available, de-identified data from the National Health and Nutrition Examination Survey (NHANES). All NHANES study protocols were approved by the National Center for Health Statistics Research Ethics Review Board, and written informed consent was obtained from all participants at the time of data collection. The present analysis did not involve direct interaction with human subjects and posed no additional risk to participants.

## Acknowledgments

The authors thank the participants and staff of the National Health and Nutrition Examination Survey (NHANES) for their valuable contributions, without which this research would not have been possible. The authors also acknowledge the National Center for Health Statistics for providing access to the NHANES datasets and linked mortality files.

## Competing interests

The authors declare that they have no competing interests.

## Funding

This research did not receive any specific grant from funding agencies in the public, commercial, or not-for-profit sectors.

## Notes

### Competing Interest Statement

The authors have declared no competing interest.

### Funding Statement

This study did not receive any funding.

